# Impact of Triple-drug Mass Drug Administration on Prevalence of Antigen and Antibodies to Lymphatic Filariasis in Samoa, 2018-2019

**DOI:** 10.64898/2026.02.02.26345345

**Authors:** Harriet LS Lawford, Helen J Mayfield, Filipina Amosa-Lei Sam, Satupaitea Viali, Tito Kamu, Robert Thomsen, Colleen L Lau

## Abstract

**Background:** In 2018, Samoa was the first country to distribute nationwide triple-drug mass drug administration (MDA) for lymphatic filariasis (LF) elimination. Prevalence of filarial antigen (Ag) is the main programmatic indicator used to define elimination target thresholds; however, anti-filarial antibodies (Ab) may provide more sensitive measures of transmission compared Ag. We aimed to investigate the utility of Ag and Ab to measure the impact of one round of triple-drug MDA on LF transmission in Samoa after 7-9 months.

**Methods:** Two community-based cross-sectional serosurveys of ≥5-year-olds were conducted in 2018 (1-3 months post-MDA) and 2019 (7-9 months post-MDA) in 35 primary sampling units. Ag was detected by Alere™ Filariasis Test Strips (FTS). Multiplex bead assays (MBA) were used to detect Ab-seropositivity (*Bm14* Ab, *Wb123* Ab) using Ab-specific mean fluorescence intensity minus background (MFI-bg). Thresholds of seropositivity were determined using finite mixture models (FMM) for log-transformed MFI-bg, defined as the mean of the presumed seronegative class plus three standard deviations. Seroprevalence was adjusted for study design, age, and gender.

**Findings:** A total of 3795 participants (mean age: 20.7; 49% male) were surveyed in 2018 and 4052 (mean age: 20.4; 48% male) in 2019. Between surveys, adjusted *Bm14* Ab prevalence decreased (31.2% [95% CI=27.0-35.8] vs 26.3% [95% CI=22.5-30.4]; *p*=0.02). In 2019 vs 2018, lower odds of *Bm14* Ab-positivity (aOR=0.76 [0.64-0.90]), *Wb123* Ab-positivity (aOR=0.70 [0.55-0.89]), and dual positivity (aOR=0.68 [0.54-0.87]) were seen. Ag-positive, microfilariae (Mf)-positive participants had significantly higher mean MFI-bg for *Wb123* Ab and *Bm14* Ab compared to Ag-positive, Mf-negative participants.

**Conclusion:** Significant reductions in *Bm14* Ab seroprevalence suggest reduced LF transmission following one round of triple-drug MDA. Thus Abs may provide a more sensitive indicator of change compared to Ag. High MFI values could help identify persistent transmission in the absence of Ag testing.

**Author summary:** Lymphatic filariasis is a mosquito-borne disease that can cause lifelong disability and stigma. The main global strategy to eliminate this disease is to treat entire at-risk communities with preventive medicines, aiming to stop transmission altogether. Progress is usually measured using rapid tests that detect parasite antigens in blood. However, as infection becomes rarer, these tests may miss remaining cases, raising concerns that ongoing transmission could go undetected.

In this study, we examined whether measuring antibodies in blood could provide earlier or more sensitive signals of infection following treatment. We analysed blood samples collected from communities in Samoa at two time points after a nationwide treatment campaign using a three-drug combination. We found clear reductions in some antibodies over time, even though antigen levels remained largely unchanged. People with active infection had much higher antibody levels than those without, suggesting antibodies may help identify areas where transmission is still occurring.

Our findings show that antibody testing can add valuable information to current monitoring approaches and may help programmes better judge whether treatment is working. This is especially important at a time when funding for disease-specific surveillance is limited, and integrated approaches are increasingly needed to protect elimination gains.

## Introduction

Lymphatic filariasis (LF), a neglected tropical disease (NTD) caused by filarial nematodes transmitted by infected mosquitoes, is a major cause of chronic disability.[1] Three species can infect humans: *Wuchereria bancrofti* (responsible for 90% of infections globally), *Brugia malayi*, and *Brugia timori*. Infected mosquitoes deposit mature third-stage filarial larvae (L3) onto a person’s skin, where they enter the host body, migrate to and nest in the lymphatic vessels, and develop into adult worms that can cause considerable damage to the lymphatic system. Chronic infection can lead to severe lymphedema and scrotal hydrocele, which often result in chronic disability, social stigma and discrimination, adverse mental health outcomes, and loss of income.[2]

In 1997, the 50th World Health Assembly resolved to eliminate LF as a public health problem, leading to the inception of the Global Programme to Eliminate LF (GPELF) in 2000 to support the global elimination of LF as a public health problem by 2020.[3] The main strategies of GPELF are two-fold; the first is to interrupt LF transmission by implementing mass drug administration (MDA) with anthelmintics to the entire at-risk population in the 72 endemic countries. MDA is effective in clearing microfilariae (Mf) in the bloodstream, thereby breaking the transmission cycle and preventing further transmission.[4] The second strategy is to alleviate suffering in the affected population through morbidity management and disability prevention.

The *World Health Organization (WHO) Roadmap for Neglected Tropical Diseases 2021-2030* (The Roadmap) was released in 2021, providing recommendations to strengthen programmatic responses to NTDs through shared goals and disease specific targets.[5] The Roadmap continues to advocate for GPELF’s efforts and aims for 58 of the 72 LF-endemic countries to have eliminated LF as a public health problem by 2030. We are now approximately halfway through The Roadmap and, to date, over 8.6 billion anthelmintic treatments have been delivered through MDA programmes, resulting in a 74% decrease in LF infections globally and 21 endemic countries validated as eliminated LF as a public health problem.[6]

Prevalence of filarial antigen (Ag) is the main indicator used in programmatic surveys to assess progress towards LF elimination, with WHO defining target Ag thresholds to determine whether LF has been eliminated as a public health problem. These thresholds vary by the causative filarial species and mosquito vector;[7] in areas where *Wuchereria bancrofti* is transmitted by *Aedes* mosquitoes, elimination is defined as an Ag prevalence of <1% measured through transmission assessment surveys (TAS), usually school-based surveys among children aged 6-7 years.[7] However, as infection prevalence declines, reliance on a fixed threshold increases the risk that residual infections remain undetected by routine rapid Ag testing, raising concerns about whether current thresholds reliably reflect true interruption of transmission.[8, 9] Anti-filarial antibodies (Ab) are generally detected at higher prevalence than Ag[10] and may appear before the development of patent infection or antigenemia.[11] Ab may therefore provide more sensitive indicators of early or low-level LF infection, enabling earlier detection of ongoing transmission. Incorporating Ab surveillance could therefore strengthen elimination efforts by supporting timely programmatic responses and reducing the risk of resurgence or premature cessation of MDA.

Ab surveillance is an emerging tool for LF monitoring. Although Ab testing is available for both *Brugia* spp. and *Wuchereria bancrofti*, current WHO guidance recommends their programmatic use only for *Brugia* spp., largely due to the widespread availability of rapid Ag tests and the inability of Ab assays to reliably distinguish between current and past *Wuchereria bancrofti* infection.[12] Nonetheless, research has demonstrated the added value of Ab testing alongside Ag surveillance.[13] A longitudinal study in American Samoa found discordant Ag and Ab patterns across three school-based TAS, with declines in Ag and *Bm14* Ab prevalence but increases in *Wb123* Ab prevalence.[14] Notably, schools with higher *Wb123* Ab prevalence in TAS-2 had significantly higher odds of Ag-positivity in TAS-3, suggesting that *Wb123* Ab may provide an earlier signal of resurgence.[14]

Multiplex bead assays (MBA) quantify Ab responses to pathogen-specific Ag using median fluorescence intensity minus background (MFI-bg) with Ab seropositivity defined by Ag-specific cut-offs.[15] MBAs enable simultaneous measurement of Ab to multiple Ags using a single sample, providing a cost-effective and resource-efficient platform for multi-disease surveillance.[15] When incorporated into a single survey framework, MBAs can reduce surveillance costs and support integrated disease monitoring approaches, as encouraged by WHO.

Samoa remains endemic for LF despite many decades of elimination efforts. Between 1965 and 1997, ten rounds of MDA of diethylcarbamazine (DEC) were distributed, with a further eight rounds of DEC and albendazole distributed under the Pacific Programme to Eliminate LF (PacELF) between 1999 and 2011.[16] In 2013, a school-based TAS in 6-7-year-olds identified ongoing transmission in Northwest Upolu (NWU), resulting in two subnational MDA rounds in this region in 2015 and 2017. All evaluation units (EUs) failed to meet elimination targets in a repeat TAS in 2017.[16]

In 2017, WHO released new guidelines recommending annual rounds of triple-drug MDA (ivermectin, diethylcarbamazine, albendazole [IDA]) for countries that were non-endemic for onchocerciasis and had experienced suboptimal results with previous rounds of two-drug regimens.[17] Subsequently, from 14-26 August 2018, Samoa was the first country in the world to implement nationwide triple-drug MDA using IDA with reported coverage of 80.2% of the total population.[18] A second round of triple-drug MDA was planned for mid-2019, but was delayed to 16-24 September 2023 due to a severe measles outbreak and the COVID-19 pandemic.[19, 20]

The Surveillance and Monitoring to Eliminate LF and Scabies from Samoa (SaMELFS) project is an ongoing operational research program to monitor and evaluate the effectiveness of triple-drug MDA on LF transmission.[16] The SaMELFS baseline survey took place from 26 September to 9 November 2018, within 1-3 months of completion of the first national distribution of triple-drug MDA. The first SaMELFS follow-up survey took place from 28 March to 17 May 2019, 7-9 months after the completion of the first round of MDA.[21] Previous SaMELFS publications from 2018 and 2019 reported no change in overall prevalence of Ag between 2018 and 2019,[22] . However, a significant reductions in mosquito infection prevalence (measured by molecular xenomonitoring [MX]) in the same time period suggests that there was a reduction in human infection following the MDA .[21]

The current study aimed to investigate the utility anti-filarial Ab to measure this impact of one round of triple-drug MDA on LF transmission in Samoa using complementary human-based indicators. The objectives of the study were to, between 2018 and 2019, (i) compare changes in seropositivity to *Bm14* Ab, *Wb123* Ab, and both *Bm14* Ab and *Wb123* Ab (dual positivity); (ii) compare changes in mean MFI-bg (mMFI-bg) of *Bm14* Ab and *Wb123* Ab; and (iii) identify and compare risk factors for Ag and Ab positivity.

## Methods

### Study setting

Samoa is a tropical archipelago in the South Pacific made up of four inhabited islands divided into four census regions: Apia Urban Area (AUA), NWU, Rest of Upolu (ROU), and Savai’i (SAV). The 2022 census estimated a total population of 205,557,[23] the majority of whom live on the islands of Upolu and SAV. *Wuchereria bancrofti* is the only species of filarial worm known to cause LF in Samoa and infection is transmitted by *Aedes* species, predominantly *Aedes polynesiensis*.[16]

### Ethics statement

Ethics approvals were granted by the Samoa Ministry of Health (MOH) and The Australian National University Human Research Ethics Committee (protocol 2018/341) and ratified by The University of Queensland’s Human Research Ethics Committee (protocol 2021/HE000895). The study was conducted in close collaboration with the Samoa MOH, the WHO country office in Samoa, and the Samoa Red Cross. Permission was obtained from village leaders prior to the commencement of the survey in their community. Written informed consent was obtained from adult participants, and verbal assent was obtained from participants aged <18 years with formal written consent obtained from a parent or guardian.

### Data source

The SaMELFS survey design has been described in detail elsewhere.[16] Briefly, both the 2018 and 2019 community-based cross-sectional serosurveys included participants ≥5 years old recruited from 35 primary sampling units (PSUs). The same 35 PSUs, each consisting of one or two villages, were sampled in both surveys and in approximately the same order, with at least a five month gap between the two surveys.[20] Thirty PSUs were randomly selected and five were purposively selected by the MOH as suspected LF hotspots based on local knowledge and high Ag prevalence from previous surveys (Figure 1).[24] To achieve the target sample size of 2000 participants aged 5-9 years and 2000 participants aged ≥10 years, a convenience survey of 5-9-year-old children was conducted in each PSU, whereby children were asked to gather at a central location for enrolment in the study.[16] Each participant completed a questionnaire collecting demographic information and participation in previous MDA programs, and had a finger prick sample of up to 400μL of blood collected into heparin microtainers. Ag was detected using Alere™ Filariasis Test Strips (FTS) (Abbott, Scarborough, ME), and two stained thick blood smears of 60 μL each were prepared from Ag-positive samples to detect Mf. Dried blood spots (DBS) were prepared for all participants (irrespective of Ag status) using Cellabs Tropbio Filter Paper Blood Collection Disks™ (six 10μL spots per person).

**Figure 1:**
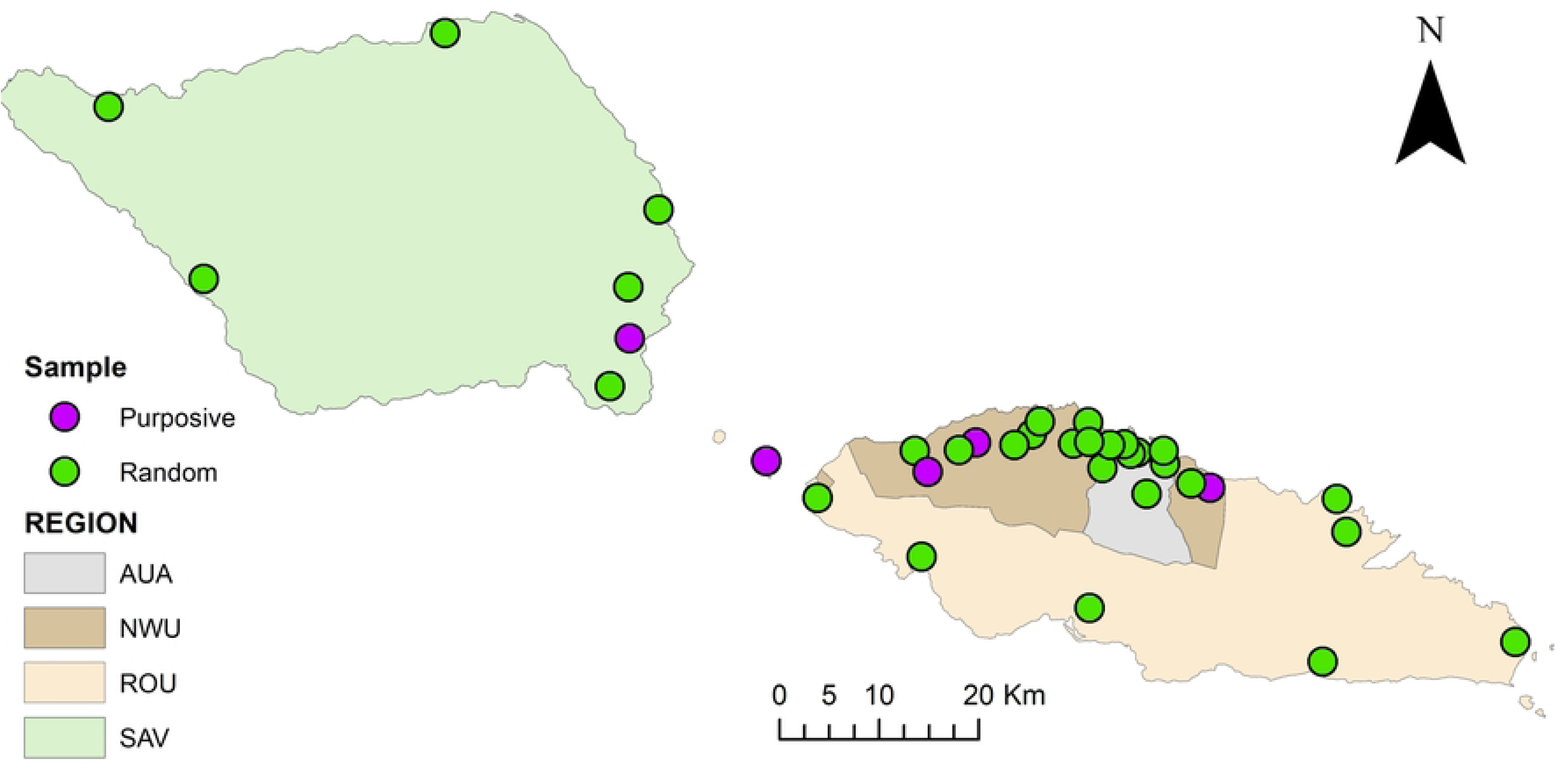
SaMELFS Samoa 2018 and 2019 surveys: Map showing administrative regions and locations of 35 PSUs (purposively selected [purple dots] and randomly selected [green dots]). *AUA = Apia Urban Area, NWU = Northwest Upolu, ROU = Rest of Upolu, SAV = Savai’i*.

### Multiplex bead assays and finite mixture modelling

DBS were sent to the Division of Parasitic Diseases and Malaria, US Centers for Disease Control and Prevention (US CDC), Atlanta, for analysis, with all Ag produced in-house. DBS were eluted into 96-well plates and then diluted to a final concentration of 1:400. All samples were tested using multiplex bead assays (MBA) for anti-filarial Ab to *Bm14* and *Wb123*. Sample plates were read on a Bio-Plex 200 instrument (Bio-Rad, Hercules, CA). For internal quality control purposes, four controls were used for the MBA analyses: a buffer blank containing the assay buffer only (in order to subtract any background noise), two pools of reference sera from known Ab-positives, and finally a negative control serum with known negative LF status. Of note, two separate bead batches were used in 2019.

A two-component Gaussian finite mixture model (FMM) was fitted separately by Ag, year, and batch to model the distribution of Ab responses from two latent subpopulations, representing presumed seronegative and seropositive individuals. Class-specific means and variances were extracted using “*lcmean*” and “*summarize*”. The class with the lower mean was assumed to represent the seronegative population. The mean plus three standard deviations of the log-transformed MFI-bg (log MFI-bg) were used to determine Ag-specific cut-offs by year and batch (see Supplementary File 1 for FMM results and Ag-specific thresholds for seropositivity). Participant were considered ‘dual positive’ if they were seropositive to both *Bm14* Ab and *Wb123* Ab.

### Statistical analysis

Data were analysed using Stata (StataCorp, Version 19.5, College Station, TX). Summary statistics with 95% confidence intervals (95% CI) for prevalence were calculated for age groups (5-9 years old vs ≥10 years old), sex (male or female), PSU selection (randomly selected vs purposively selected), and region (AUA, NWU, ROU, and SAV).

Prevalence estimates were adjusted for selection probability and clustering using methods that have been described previously.[16] Briefly, weighting for selection probability and clustering were based on the 2016 Samoa Census[25] and performed using the “*svyset*” command in Stata with PSU as the unit of clustering. Age group and gender standardised weights were applied using information from the 2016 Samoa Census.[25] Prevalence estimates for the two age groups were adjusted for selection probability and clustering, and standardised for gender but not age. Prevalence estimates for all ages ≥5 years were adjusted for selection probability and clustering and standardised for sex (except when comparing between sex) and age using 5-year age bands.

Seroprevalence, defined as the percent of individuals with MFI-bg above the threshold determined by FMM, was calculated using the “*proportion*” command for the total population and stratified by age (5–9-years-old vs ≥10 years-old) and PSU selection (randomly vs purposively-selected). For each Ab, the mean population log MFI-bg value was calculated for each year and batch, and the 95% CI were calculated assuming a normal distribution of the log10 MFI-bg values. Change in *Bm14* Ab-positivity, *Wb123* Ab-positivity, and seropositivity to both *Bm14* Ab and *Wb123* Ab between 2018 and 2019 were compared between participants testing i) Ag-positive (inclusive of Mf-positives and Mf-negatives), ii) Ag-positive and Mf-positive; iii) Ag-positive and Mf-negative, and iv) Ag-negative Changes in prevalence and mean MFI-bg between years were tested for statistical significance using Wald tests with Stata’s “*lincom*” command.

Logistic regression was used to assess associations between demographic and behavioural data (taken from the survey questionnaire) and Ab positivity. Variables with *p*<0.2 on univariable analyses were tested using backward stepwise multivariable logistic regression; variables were sequentially removed from the multivariable models to arrive at the most parsimonious models, in which variables with *p*<0.05 were retained.

## Results

### Participant characteristics

In total, 4420 and 4493 participants were recruited in 2018 and 2019, respectively. Following exclusion of participants aged <5 years (480 in 2018 and 201 in 2019 who were recruited for a scabies survey only and did not have a blood sample collected), participants with no MBA results (89 in 2018 and 231 in 2019), and those with invalid FTS results (56 in 2018 and nine in 2019), a total of 3795 were included from 2018 and 4052 from 2019 surveys (Supplementary Figure 1). Demographic characteristics of participants recruited in 2018 and 2019 were similar (Table 1).

**Table 1:** Demographic characteristics of recruited participants in 2018 and 2019, Samoa.

|  | 2018 | 2019 | *p*-value* |
| --- | --- | --- | --- |
| **Total participants recruited (N)** | **3795** | **4052** |  |
| **Mean participant age in years (range)** | 20.7 (5-90) | 20.4 (5-89) | 0.56 |
| **Total male participants (%)** | 1853 (49%) | 1934 (48%) | 0.33 |
| **Total recruited via convenience survey (%)** | 1512 (40%) | 1560 (39%) | 0.56 |
| **Total participants from purposive PSUs (%)** | 518 (14%) | 618 (15%) | **0.04** |

PSU = primary sampling unit. \*Statistical differences tested using Student's *t*-test or Pearson's $\chi^2$ Test

### Crude antigen, microfilariae, and antibody prevalence

Overall, 7,847 participants from the 35 PSUs had both Ag and Ab results; 117 (3.1%; 95% confidence interval [95% CI] =2.6-3.7) and 137 (3.4%; 95% CI=2.9-4.0) participants tested Ag-positive in 2018 and 2019, respectively (*p*=0.46). Of these participants, 15/117 (x%) and 31/137 (x%) were Mf-positive. Assuming that Ag-negative participants were also Mf-negative, the overall Mf prevalence increased significantly between 2018 and 2019 from 0.4% (95% CI=0.2-0.7) to 0.8 (95% CI=0.5-1.1) (*p*=0.03). There was a significant decrease in *Bm14* Ab (from 24.1% [95% CI=22.8-25.5; n=916] in 2018 to 20.3% [95% CI=19.1-21.6; n=823] in 2019) (*p*<0.001), and *Wb123* Ab prevalence (from 8.0% [95% CI=7.1-8.9; n=302) in 2018 to 6.2% (95% CI=5.5-7.0; n=252) in 2019 (*p*=0.003). Overall, individuals dual positive for both *Bm14* Ab and *Wb123* Ab decreased significantly from 7.4% (95% CI=6.6-8.3; n=280) in 2018 to 5.6% (95% CI=4.9-6.3; n=225) in 2019 (*p*=0.001).

### Adjusted antigen, microfilariae, and antibody prevalence

Following adjustment, there was no change in Ag prevalence (3.7% [95% CI=2.6-5.2] vs 4.6% [95% CI=3.3-6.6]; *p*=0.38), adjusted Mf prevalence (0.6 [95% CI=0.3-1.2] vs 0.9 [95% CI=0.5-1.7]; *p*=0.32), adjusted *Wb123* Ab prevalence (11.4% [95% CI=8.7-14.8] vs 8.6% [95% CI=6.7-10.9]; *p*=0.052), or adjusted seropositivity for both *Bm14* Ab and *Wb123* Ab (10.5% [95% CI=8.1-13.5] vs 7.9% [6.2-10.1]; *p*=0.06). A significant decrease was seen in adjusted *Bm14* Ab prevalence (31.2% [95% CI=27.0-35.8] vs 26.3% [95% CI=22.5-30.4]; *p*=0.02) (Figure 2).

**Figure 2:**
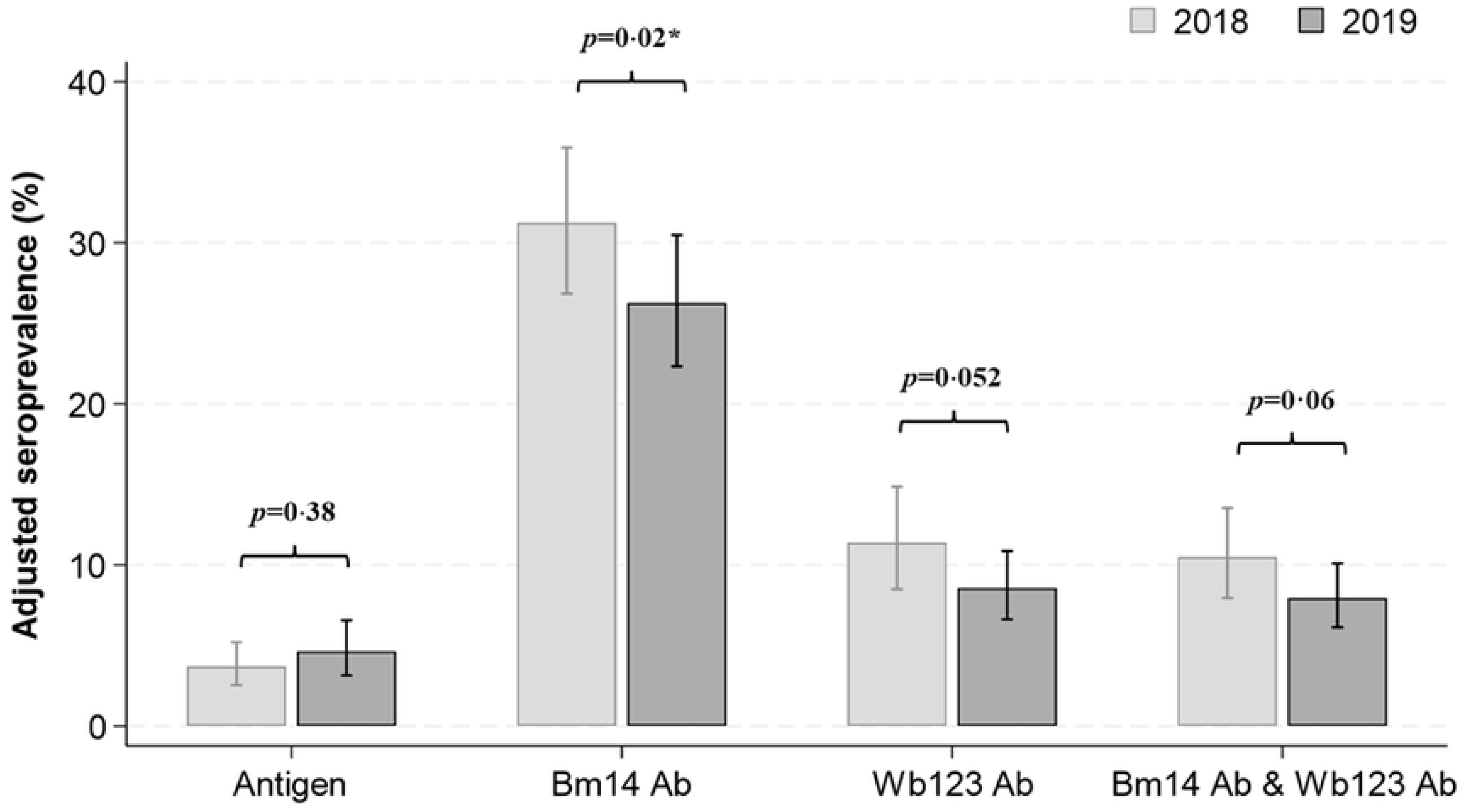
Prevalence of lymphatic filariasis antigen and antibodies (*Bm14* and *Wb123*) in 2018 (light grey) and 2019 (dark grey), Samoa. Adjusted for sampling design and standardised by age and sex. *\*Significant differences in prevalence between years assessed using Wald’s test*.

### Adjusted antigen, microfilariae, and antibody prevalence by age and sex

When stratified by age group (5-9 years and ≥10 years), no change was seen in Ag prevalence (5-9 years: 1.3% vs 1.3% [*p*=0.96]; ≥10 years: 4.3% vs 4.7% [*p*=0.80]), *Wb123* Ab prevalence (5-9 years: 2.3% vs 2.0% [*p*=0.54]; ≥10 years: 12.8% vs 9.3% [*p*=0.08]) or the proportion of individuals testing dual positive (5-9 years: 2.1% vs 1.7% [*p*=0.44]; ≥10 years: 11.9% vs 8.5% [*p*=0.054]) between 2018 and 2019. However, significant decreases were seen in *Bm14* Ab prevalence for ≥10-year-olds between 2018 and 2019 (5-9 years: 12.0% vs 10.5% [*p*=0.41]; ≥10 years: 34.2% vs 27.9% [*p*-value: 0.03]) (Figure 3 and Supplementary Table 1).

**Figure 3:**
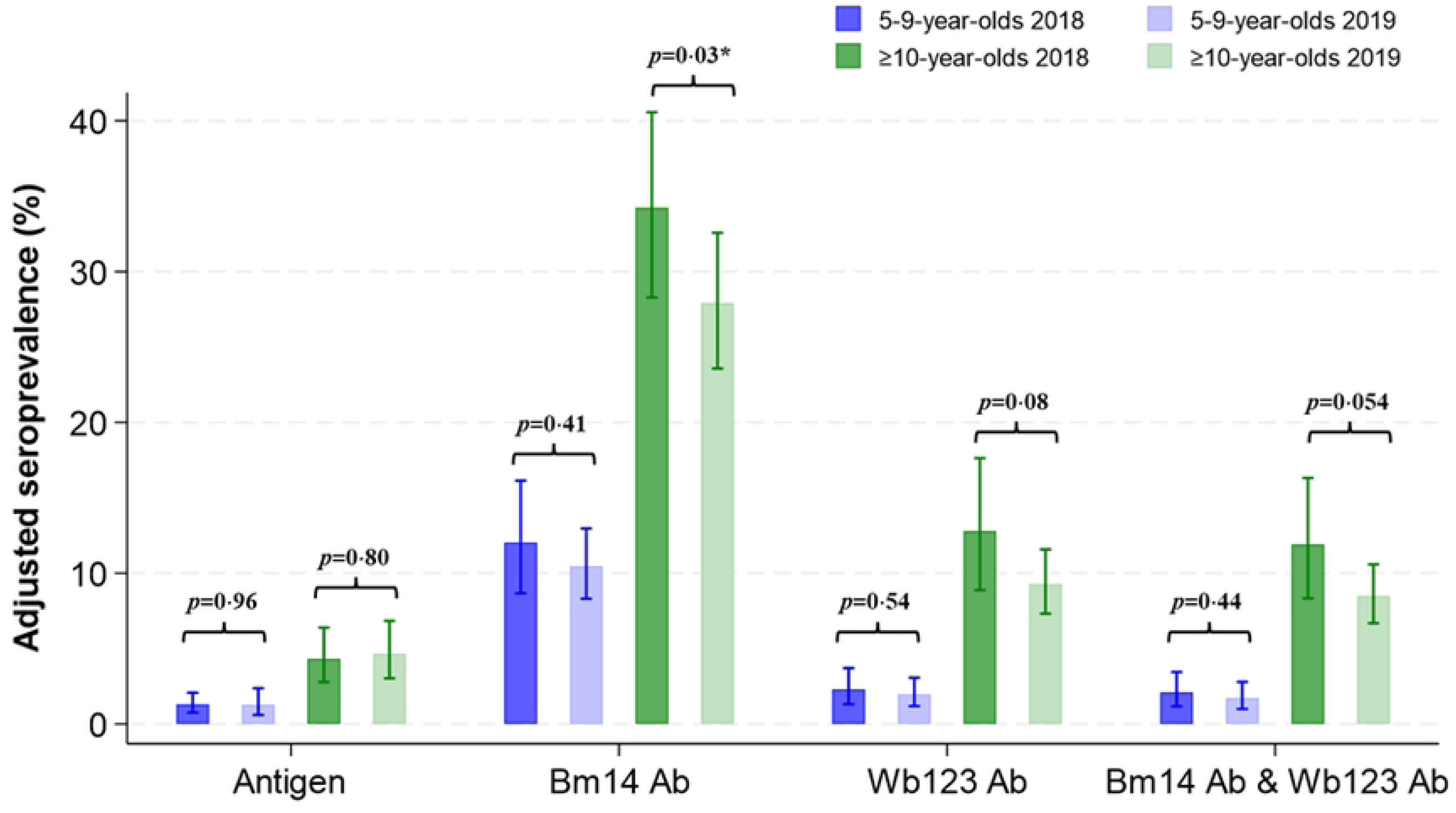
Prevalence of lymphatic filariasis antigen and antibodies (*Bm14* and *Wb123*) in 2018 (dark shading) and 2019 (light shading) among 5-9-year-olds (blue bars) and ≥10-year-olds (green bars), Samoa. Adjusted for sampling design and standardised by sex. *\*Significant difference in prevalence between years assessed using Wald’s test*.

When stratified by sex, females had a significant decrease in *Bm14* Ab prevalence (from 24.1% [95% CI=19.9-28.8] to 18.6% [95% CI=15.1-22.6]; *p*=0.03) *Wb123* Ab prevalence (from 8.4% [95% CI=5.9-12.0] to 4.1% [95% CI=2.9-5.9]; *p*=0.01), and dual seropositivity (from 7.8% [95% CI=5.3-11.4] to 3.5% [95% CI=2.4-5.2]; *p*=0.01) between years. There was no significant change in Ag prevalence or Mf prevalence. For male participants, there was no change in prevalence between years for any seromarker (Figure 4 and Supplementary Table 1).

**Figure 4:**
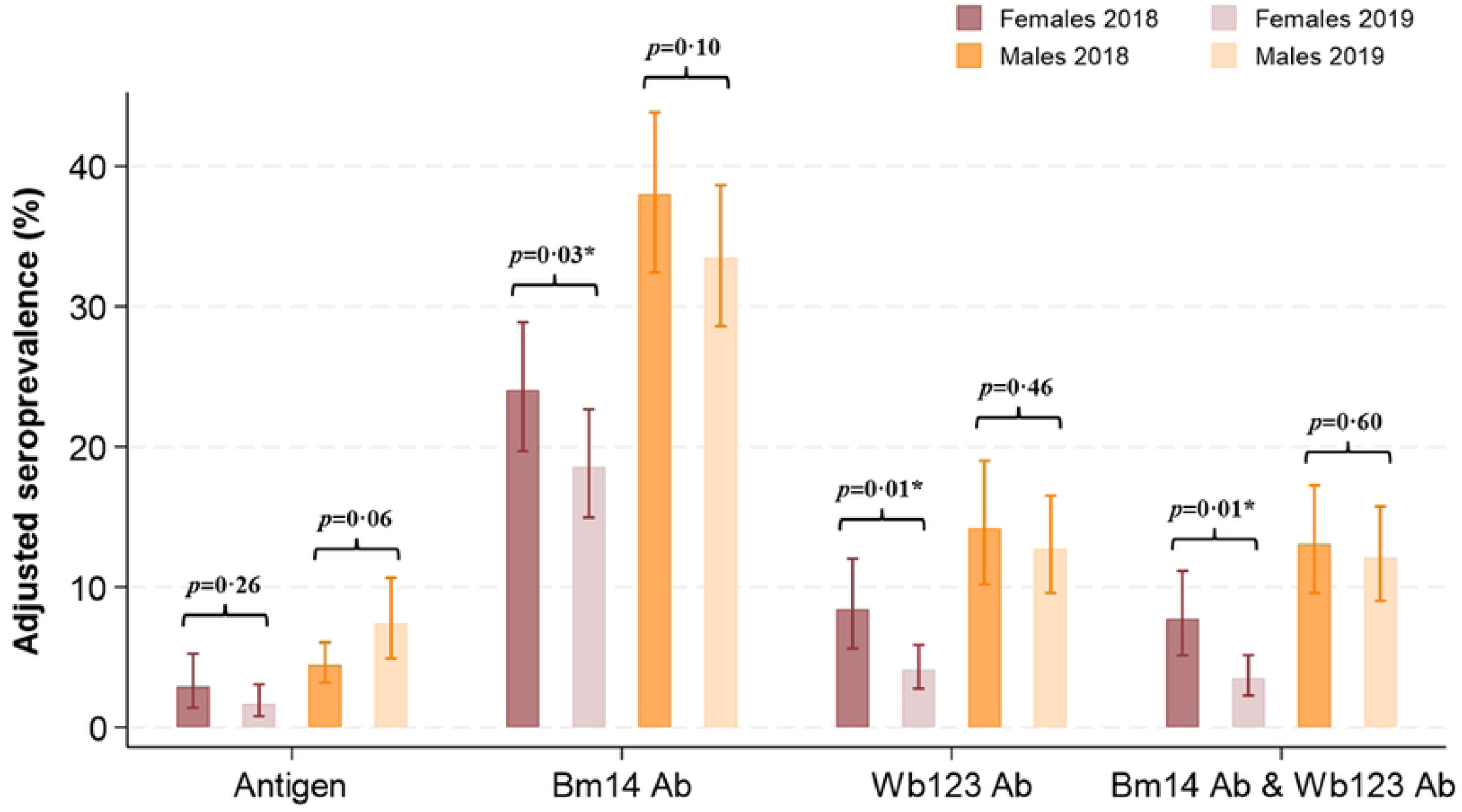
Prevalence of lymphatic filariasis antigen and antibodies (*Bm14* and *Wb123*) in 2018 (dark shading) and 2019 (light shading) among females (pink bars) and males (orange bars), Samoa. Adjusted for sampling design and standardised by age. *\*Significant difference in prevalence between years assessed using Wald’s test*.

### MFI-bg values for *Bm14* Ab and *Wb123* Ab by age group and sex

Mean log10 MFI-bg values (mMFI) for *Bm14* Ab and *Wb123* Ab in 2018 and 2019 are shown in Figure 5 and Supplementary Table 2. No change was seen in mMFI for *Bm14* Ab between 2018 and 2019 (1.81 [95% CI=1.69-1.93] vs 1.92 [95% CI=1.82-2.01]; *p*=0.11). However, an increase in mMFI for *Wb123* Ab was seen (2.20 [95% CI=2.12-2.29] vs 2.39 [95% CI=2.33-2.45]; *p*<0.001). When stratified by age and sex, mMFI for *Bm14* Ab increased between 2018 and 2019 among 5-9-year-olds only (1.29 [95% CI=1.20-1.39] vs 1.50 [95% CI=1.43-1.56]; *p*<0.001). For *Wb123* Ab, increases in mMFI were seen for all subgroups: 5-9-year-olds (1.92 [95% CI=1.84-2.00] vs 2.12 [95% CI=2.08-2.17]; *p*<0.001), ≥10-year-olds (2.25 [95% CI=2.14-2.36] vs 2.42 [95% CI=2.35-2.49]; *p*=0.01), males (2.30 [95% CI=2.20-2.39] vs 2.49 [95% CI=2.42-2.57]; *p*<0.001), and females (2.10 [95% CI=2.01-2.19] vs 2.28 [95% CI=2.23-2.34]; *p*=0.003) (Figure 6).

**Figure 5:**
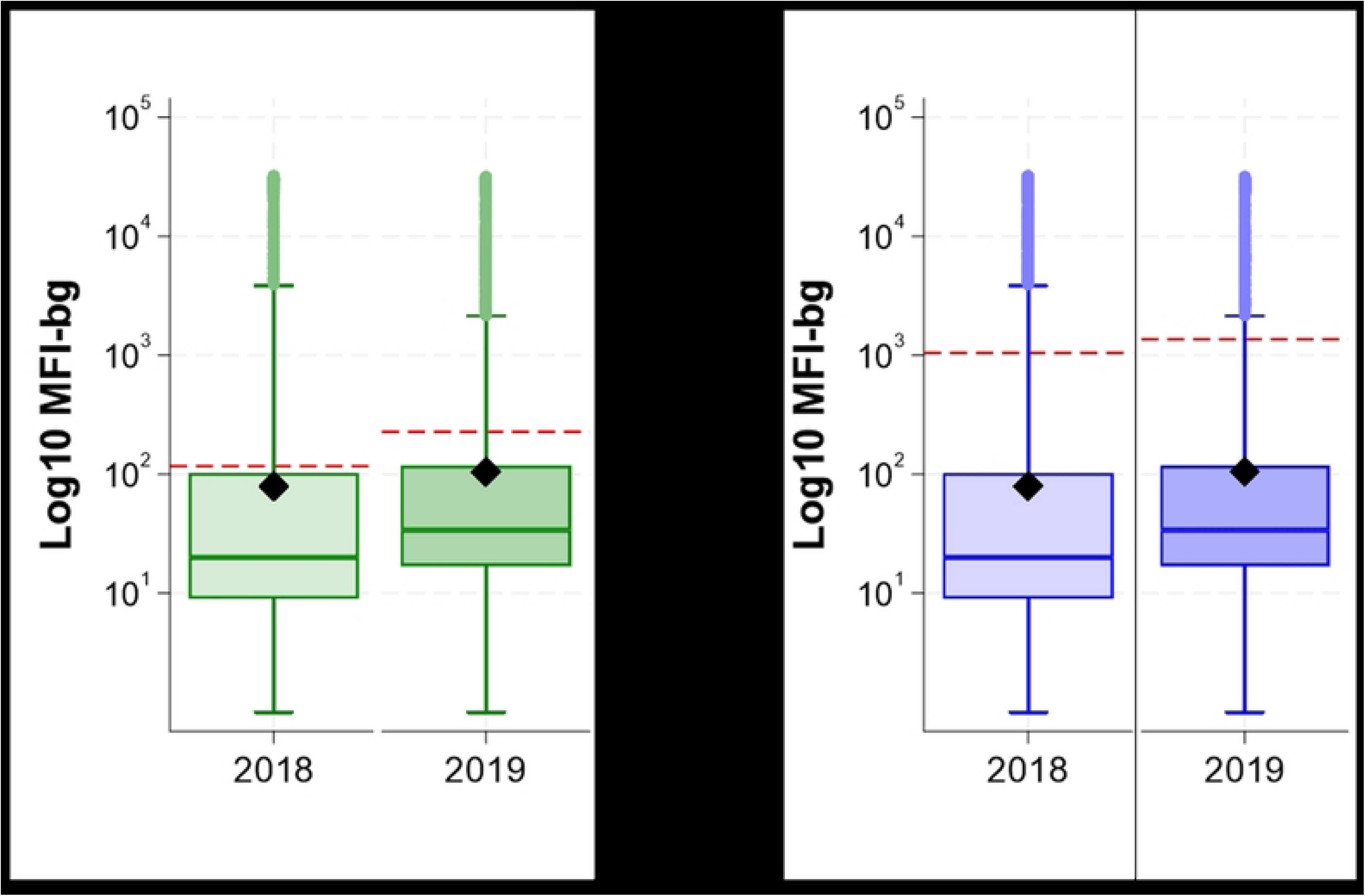
Mean log10 MFI-bg values for *Bm14* antibody (green) and *Wb123* antibody (blue) for lymphatic filariasis in 2018 and 2019, Samoa. Red line indicates antibody-specific cut-off values for positivity. Centre line indicates median, and the box represents the 25^th^ and 75^th^ centiles. Black diamonds indicate mean log10 MFI-bg values, adjusted for survey design and standardised for age and sex. Whiskers represent outliers. Log10 MFI-bg values were analysed on the log10 scale due to a skewed distribution.

**Figure 6:**
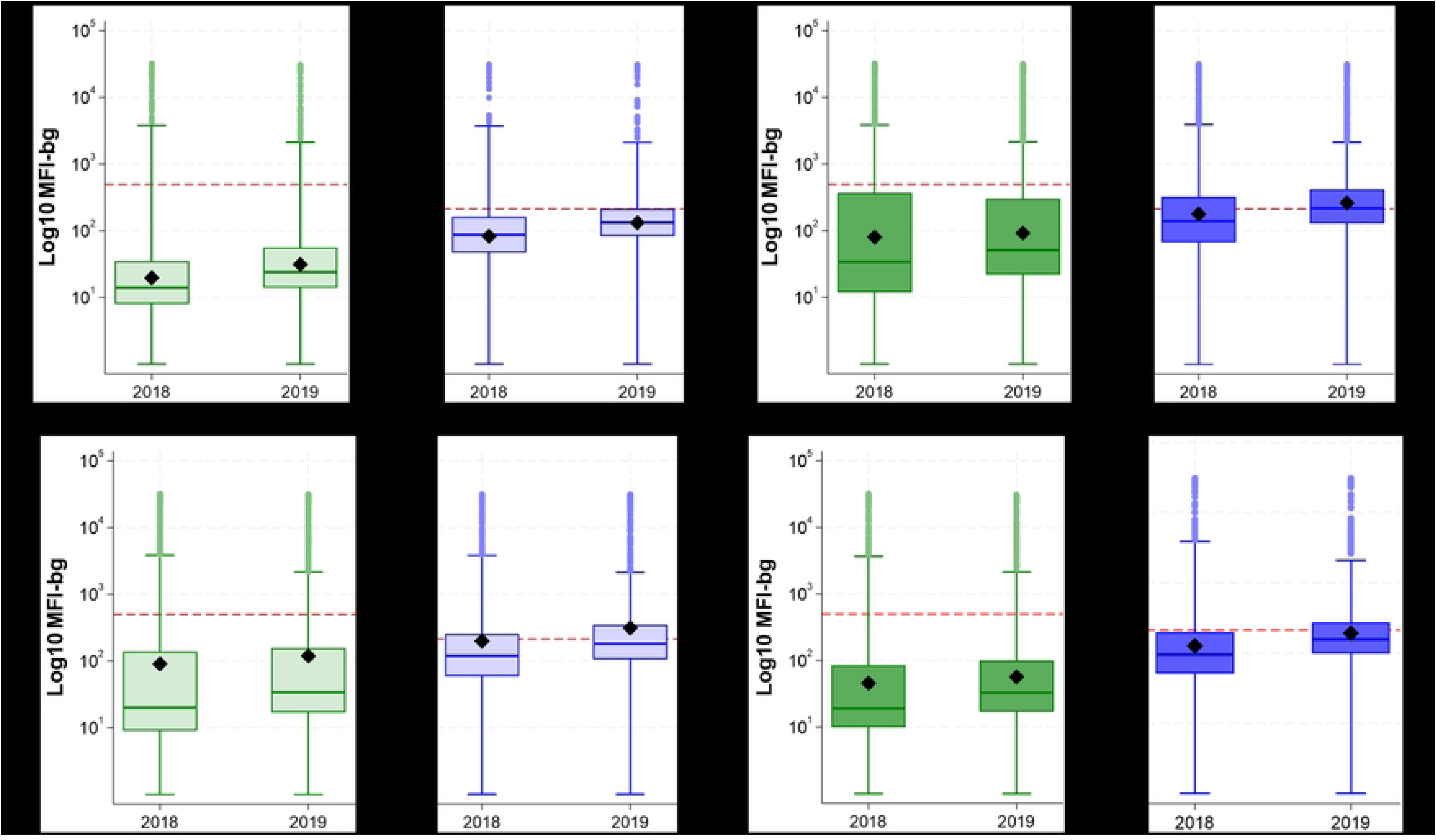
Mean log10 MFI-bg values of *Bm14* antibody (green) and *Wb123* antibody (blue) for lymphatic filariasis by age group (adjusted for survey design and standardised for sex) and sex (adjusted for survey design and standardised for age) in 2018 and 2019, Samoa. Red line indicates antibody-specific cut-off values for antibody positivity. For box plots, centre line indicates median, and the box represents the 25^th^ and 75^th^ centiles. Black diamonds indicate mean log10 MFI-bg values. Whiskers represent outliers. *Significant differences in mean log10 MFI-bg between years assessed using adjusted Wald test*.

### Serostatus and MFI-bg values for *Bm14* Ab and *Wb123* Ab by Ag-positivity and Mf-positivity

Change in seroprevalence to *Bm14* Ab, *Wb123* Ab, and dual positivity between 2018 and 2019 are shown in Table 2. Among all Ag-positive participants (inclusive of both Mf-positives and Mf-negatives), there was a significant decrease in seroprevalence for *Bm14* Ab (92.8% vs 77.5%; *p*=0.001). Among participants testing Ag-positive and Mf-positive, there was a significant increase in seroprevalence for *Bm14* Ab (91.9% to 99.0%; *p*=0.01), *Wb123* Ab (91.9% to 99.0%; *p*=0.01), and those testing dual positive (91.9% to 99.0%; *p*=0.01) from 2018 to 2019. Among participants testing Ag-positive but Mf-negative, there was a significant decrease in the proportion testing positive for *Bm14* Ab (92.6% to 76.3%; *p*<0.001) and dual positive (83.4% to 72.7%; *p*=0.03). Lastly, among Ag-negative participants, there was a significant increase the proportion testing *Wb123* Ab positive (29.9% to 41.5%; *p*=0.005).

**Table 2:**
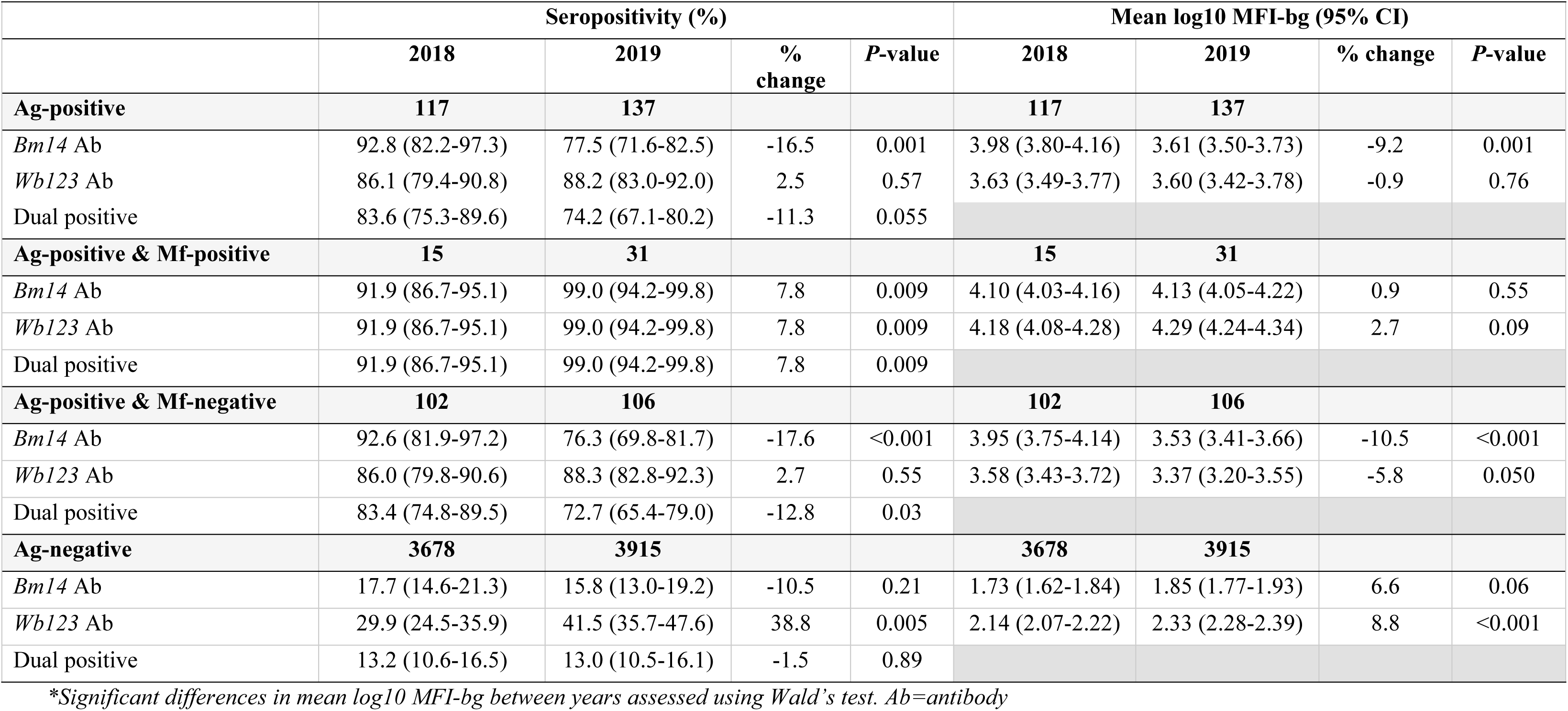
Difference between years for seropositivity and mean log10 Median Fluorescent Intensity-background (MFI-bg) values for *Bm14* antibody and *Wb123* antibody for lymphatic filariasis by antigen and microfilaria status, in 2018 and 2019, Samoa.

Changes in mMFI-bg results by participant serostatus are presented in Figure 7 and Supplementary Table 3. A significant decrease in *Bm14* Ab mMFI-bg was seen (3.98 vs 3.61; *p*=0.001) from 2018 to 2019 among all Ag-positive participants, with no change in *Wb123* Ab mMFI-bg. mMFI-bg declined significantly for participants testing Ag-positive and Mf-negative for both Ab (*Bm14* Ab: 3.95 to 3.53; *p*<0.001 and *Wb123* Ab: 3.58 to 3.37; *p*=0.050). An increase in mMFI-bg was seen between years for Ag-negative participants, significantly so for *Wb123* Ab mMFI-bg (2.14 to 2.33; *p*<0.001).

**Figure 7:**
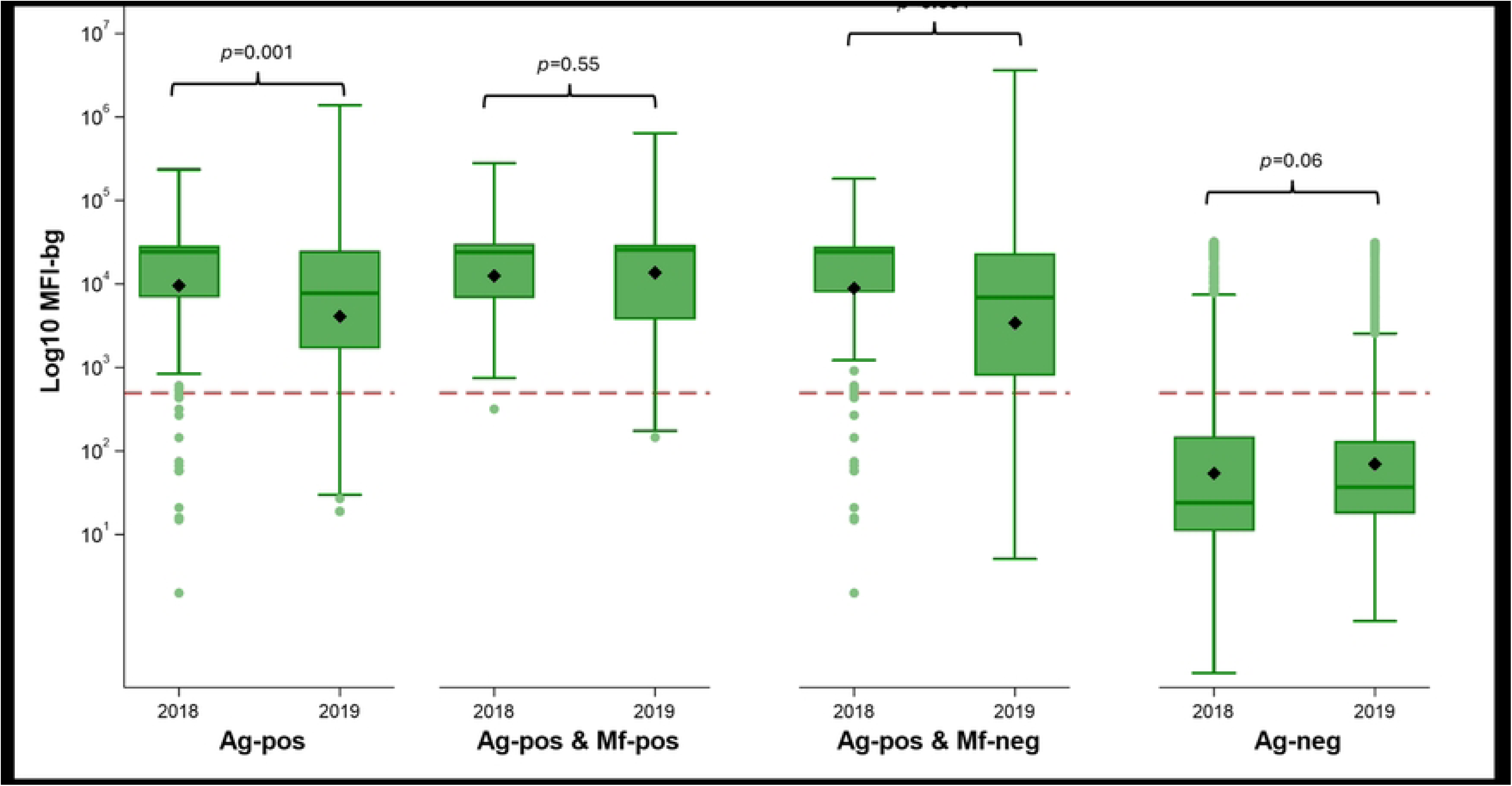
Mean log10 MFI-bg values of *Bm14* antibody (green) and *Wb123* antibody (blue) for lymphatic filariasis by antigen and microfilariae status (adjusted for survey design and standardised for age and sex) in 2018 and 2019, Samoa. Red line indicates antibody-specific cut-off values for antibody positivity. Centre line indicates median, and the box represents the 25^th^ and 75^th^ centiles. Black diamonds indicate mean log10 MFI-bg values. Whiskers represent outliers. *Significant differences in mean log10 MFI-bg by status assessed using adjusted Wald test*.

In 2019, *Bm14* Ab mMFI-bg remained significantly higher among Ag-positive individuals compared with Ag-negative individuals; however, unlike in 2018, *Bm14* Ab mMFI-bg were significantly higher among those with concurrent Ag and Mf positivity than among Ag-positive but Mf-negative individuals (mean difference +0.5%; *p*=0.03). In both 2018 and 2019, *Wb123* Ab mMFI-bg were significantly higher among Ag-positive individuals compared with Ag-negative individuals, with significantly higher mMFI-bg observed among those with concurrent Ag and Mf positivity compared to participants who were Ag-negative (*p<*0.001) and Ag-positive but Mf-negative (2018: *p*=0.03; 2019: *p<*0.001) (Supplementary Table 3 and Supplementary Figure 2).

### Differences in risk factors for positivity between 2018 and 2019

Similar risk factors for seropositivity were seen in 2018 and 2019. For all seromarkers in both years, significant associations were seen between positivity and older age (³10-year-olds vs 5-9-year-olds), males (vs females), and residing in purposively selected PSUs (vs randomly selected PSUs). Only regional associations changed between 2018 and 2019. No association was seen between region of residence and seropositivity in 2018 (except for *Bm14* Ab-positivity where residents of NWU had 2.27 [95% CI=1.57-3.30] the odds of positivity compared to residents of AUA). However, in 2019, increased odds of positivity were seen for all seromarkers in all regions compared to AUA (except SAV) in 2019 (Supplementary Table 4 and Supplementary Figure 3).

The adjusted odds of participants testing positive to Ag and Abs are shown in Figure 8 and Supplementary Table 5. Compared to 2018, the odds of testing Ag-positive in 2019 were significantly higher among residents of purposively selected PSUs (adjusted odds ratio [aOR]=1.54 [95% CI=1.08-2.20]), residents of SAV (aOR=1.76 [95% CI=1.34-2.30]), and lower among residents of AUA (aOR=0.28 [95% CI=0.10-0.78]). The odds of testing *Bm14* Ab-positive were significantly lower in 2019 compared to 2018 for the overall population (aOR=0.76 [95% CI=0.64-0.90]; *p*=0.002), among residents of randomly selected PSUs (aOR=0.76 [95% CI=0.52-0.88]) residents of NWU (aOR=0.68 [95% CI=0.52-0.88]), among female participants (aOR=0.68 [95% CI=0.52-0.88]), and among participants aged ≥10 years (aOR=0.75 [95% CI=0.60-0.94]). Similarly, the odds of *Wb123* Ab-positivity were lower for the overall population (aOR=0.70 [0.55-0.89]), among participants from randomly selected PSUs (aOR=0.69 [95% CI=0.54-0.89]), among residents of NWU (aOR=0.59 [95% CI=0.42-0.84]), females (aOR=0.59 [95% CI=0.41-0.84]), and for ≥10-year-olds (aOR=0.68 [95% CI=0.51-0.89]) compared to 2018. Lastly, the odds of testing dual positive were significantly lower overall in 2019 vs 2018 (aOR=0.68 [95% CI=0.54-0.87]), among participants from randomly selected PSUs (aOR=0.67 [95% CI=0.53-0.87]), residents of NWU (aOR=0.60 [95% CI=0.42-0.85]), males (aOR=0.79 [95% CI=0.63-0.99]) and females (aOR=0.56 [95% CI=0.39-0.82]), and participants aged ≥10 years (aOR=0.66 [95% CI=0.50-0.86]).

**Figure 8.**
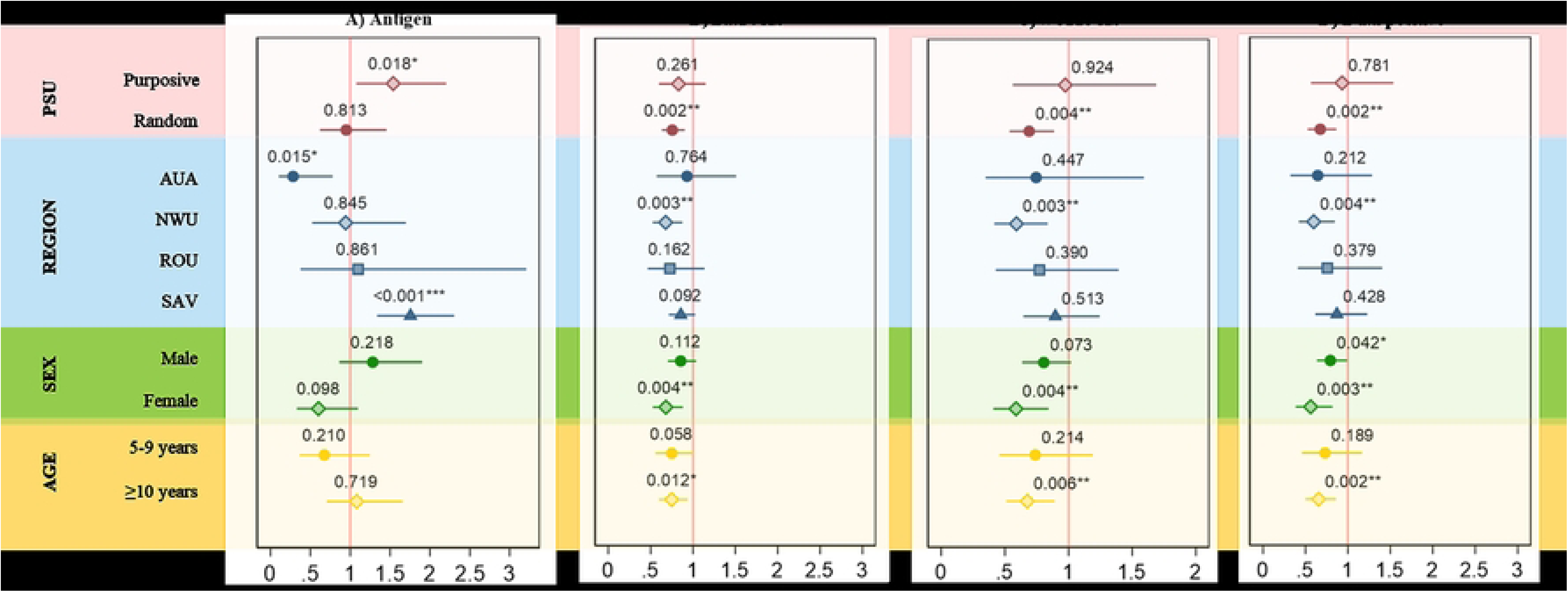
Adjusted odds ratios (and *p-*values) for associations with seropositivity to lymphatic filariasis A) antigen, B) *Bm14* antibody, C) *Wb123* antibody, and D) both *Wb123* and *Bm14* antibodies in 2019 compared to 2018, Samoa. The red solid line denotes the “line of null effect” (adjusted odds ratio =1). \**p*<0.05 \*\**p*<0.01 \*\*\**p*<0.001. *Regions: AUA (Apia Urban area) NWU (Northwest Upolu), ROU (Rest of Upolu), SAV (Savai’i); Ab=antibody; CI=Confidence Interval*.

## Discussion

Our study found that a single round of triple-drug MDA had differing effects over time on LF Ab seromarkers in Samoa. We found a significant reduction in seroprevalence to *Bm14* Ab and non-significant reductions in *Wb123* Ab and dual positivity, supported by significantly reduced odds of seropositivity for all Ab seromarkers in 2019 compared to 2018. We also observed significantly higher mMFI-bg levels among Ag-positive, Mf-positive participants compared with Ag-positive, Mf-negative participants, suggesting that elevated mMFI-bg values may help identify communities with persistent transmission. Finally, these Ab-specific changes in mMFI-bg at baseline and follow-up provide insights into the post-MDA dynamics of *Bm14* Ab and *Wb123* Ab and highlight how temporal Ab patterns can support ongoing LF monitoring.

Our finding of a significant reduction in *Bm14* Ab seroprevalence suggests a decrease in LF transmission following a single round of triple-drug MDA, consistent with other studies reporting significant reductions in mosquito infection prevalence between SaMELFS 2018 and 2019.[21] Similarly, a study from Haiti conducted within a similar time frame reported significant declines in *Bm14* Ab seroprevalence following successful MDA.[26] In The Gambia, very low *Wb123* and *Bm14* seroprevalence was interpreted as evidence of interrupted LF transmission.[27] Although we found no reduction in Ag prevalence, this may reflect ongoing or low level transmission signals that are missed by Ag-based diagnostics alone, highlighting the greater sensitivity of Ab-based measures for post-MDA surveillance.

Prior research suggests that *Bm14* Ab-positivity is associated with the presence of adult worms and/or recent exposure to infective L3,[28] and that *Bm14* Ab is highly sensitive for classifying individuals as Ag-positive, indicating that these markers reflect similar stages of infection.[10] In contrast, *Wb123* Ab is considered an early marker of infection, primarily expressed by L3,[29] and has been seen to develop months to a year before the appearance of Mf, consistent with its role as a marker of pre-patent infection.[11]

Between 2018 and 2019, we observed distinct temporal dynamics in seroprevalence and mMFI-bg for *Bm14* Ab and *Wb123* Ab. In 2018, *Bm14* Ab mMFI-bg was similarly elevated among Ag-positive individuals regardless of Mf status, consistent with the theory that *Bm14* reflects cumulative exposure.[30] Following MDA, *Bm14* Ab mMFI-bg declined significantly among Ag-positive but Mf-negative individuals, resulting in clear differentiation between those with and without Mf. This pattern suggests that the observed shift was driven by waning Ab levels among Ag-positive, Mf-negative individuals rather than increases among those with concurrent Ag and Mf positivity, suggesting that *Bm14* mMFI-bg declines following clearance of Mf.

In contrast, *Wb123* Ab mMFI-bg was significantly higher among Mf-positive, Ag-positive participants compared with Mf-negative, Ag-positive individuals in both 2018 and 2019, with increasing separation over time. Persistently high *Wb123* Ab mMFI-bg among Mf-positive individuals, and declining levels among Mf-negative individuals, support its utility as an early marker of infection or recent infection in post-intervention settings. Notably, both *Bm14* Ab and *Wb123* Ab showed significant increases in mMFI-bg among Ag-negative individuals between years, underscoring the importance of stratified and longitudinal interpretation of Ab markers in post-MDA contexts.

Our findings suggest the potential to explore MFI-bg thresholds as a means to distinguish Mf-positive (active infection) from residual Ag positivity over time. If validated, such thresholds would substantially increase the programmatic value of Ab surveillance by enabling LF programmes to more precisely identify and target foci of active LF transmission. Testing for *Wb123* Ab may be particularly well suited to threshold-based approaches, as very high *Wb123* Ab MFI-bg were seen even prior to intervention.

To date, only one other study has investigated the impact of triple-drug MDA on Ab prevalence. In this study by Supali *et al* (2023) in Indonesia, 35 Mf-positive participants were treated with IDA and followed up one year later.[31] Using an in-house indirect ELISA on 34/35 paired pre- and post-treatment blood samples, the authors found that 33/35 participants cleared Mf, but *Bm14* Ab prevalence remained almost unchanged.[31] Possibly, a longer interval between surveys may be needed to observe a true reduction in Ab seroprevalence after adult worms have been cleared from the body. Notably, Supali *et al* (2023) found one Mf-positive participant who was *Bm14* Ab-negative before and after treatment, consistent with reports that up to 10% of Mf-positive individuals may be *Bm14* Ab-negative in studies from Indonesia[32] and Haiti.[33]

An interesting finding from our study was sex-specific declines in *Bm14* Ab seroprevalence, *Wb123* Ab seroprevalence, and dual positivity among females only, highlighting heterogeneity in serological responses following MDA. Similar shifts towards higher LF prevalence among males during and after MDA has been reported elsewhere,[34] and may reflect a natural ‘epidemiological shift’ towards increasingly focal transmission concentrated within high-risk groups and areas. This pattern underscores the importance of achieving high MDA coverage among adult males and ensuring they are included in surveillance activities. Other studies have suggested that females may mount an earlier Ab response (but not Ag response),[11] which may partly explain the observed decreases in Ab seroprevalence among females in this study.

A key limitation in this study was the uncertainty around cut-offs used to determine seropositivity. FMM are commonly used to derive seropositivity thresholds; however, standardised methods are essential if Ab seroprevalence is to be used as a monitoring and evaluation tool. Studies from Vanuatu have reported reluctance among programmes to use Ab assays because *“consistent, reproducible and reliable international assay standards and cut-offs to determine positivity”* were lacking, with mixture modelling therefore used to derive robust, locally appropriate thresholds.[35] Other work warned that false positives in low-prevalence settings could lead to unnecessary interventions and has highlighted the need for larger sample sizes or lower critical cut-offs to maintain statistical power, alongside calls for WHO to standardise validation frameworks to ensure comparability across settings.[36] Uncertainty around cut-off values can risk misclassification of infection status, particularly in low-transmission settings, emphasising the need for more sensitive and specific recombinant Ags to support confident interpretation of results.[37]

Despite these knowledge gaps, there are several benefits to incorporating Ab testing alongside Ag testing for monitoring LF prevalence. Firstly, multiplex technology enable the concurrent measurement of Abs to hundreds of pathogen-specific Ag from a single sample, including dried blood spots. This cost-effective approach supports a shift from single-disease surveillance to integrated, multi-disease platforms. This is particularly critical in the current global health funding climate, where cuts to NTD-specific funding increasingly limit stand-alone surveillance, and integration into established programmes, such as serosurveys for vaccine-preventable diseases, may represent one of the few sustainable pathways for continued NTD surveillance.

Secondly, research suggests that Ab-based surveillance may require fewer individuals than Ag testing to detect transmission signals,[38] reducing the burden on communities, particularly when surveillance is targeted to suspected hotspots. Consistent with this, our study found that Ag-positive, Mf-positive participants had significantly higher mMFI for *Wb123* Ab and *Bm14* Ab compared with Ag-positive, Mf-negative participants, supporting the utility of Ab monitoring to identify ongoing transmission.

As research continues and knowledge gaps and optimise surveillance strategies and diagnostic tools for LF, rich longitudinal data that capture changes in Ag and Ab dynamics over time are highly valuable. Such data broaden our understanding of the population-level impact of triple-drug MDA and inform interpretation of LF transmission signals for programmatic action (be they based on Ag-positivity, Mf-positivity, Ab-positivity, MX results, or a combination of all). However, a key challenge limiting the utility of Ab serosurveillance for LF is the lack of field deployable point-of-care tests.

## Conclusion

Overall, our findings indicate that serological tools have the potential to support surveillance, guide programmatic decision-making, and help monitor the impact of MDA in countries aiming to interrupt LF transmission. However, our results underscore the complexity of interpreting serological markers in post-MDA settings. While Ab surveillance cannot currently distinguish between past and active infection, combining Ab and Ag testing, and examining high mMFI-bg in particular, may support the identification of foci with ongoing transmission. Continued longitudinal surveillance and improved understanding of Ab kinetics are essential to refine post-MDA LF monitoring strategies and enable timely, targeted programmatic interventions. Further longitudinal studies are therefore needed to characterise Ab kinetics following MDA and to define how Ab-based measures can be most effectively interpreted for LF surveillance and control.

## Data sharing statement

All relevant data are within the paper. We are unable to provide individual-level antigen prevalence data and demographic data because of the potential for breaching participant confidentiality. The communities in Samoa are very small, and individual-level data such as age, sex, and village of residence could potentially be used to identify specific persons. For researchers who meet the criteria for access to confidential data, the data are available on request from the Human Ethics Officer at The University of Queensland.

## Data Availability

All relevant data are within the paper. We are unable to provide individual-level prevalence data and demographic data because of the potential for breaching participant confidentiality. The communities in Samoa are very small, and individual-level data such as age, sex, and village of residence could potentially be used to identify specific persons. For researchers who meet the criteria for access to confidential data, the data are available on request from the Human Ethics Officer at The University of Queensland.

## Acknowledgements

We gratefully acknowledge the contributions of the following organisations: Samoa Ministry of Health, Samoa Red Cross, WHO Samoa Country Office, and US Centers for Disease Control and Prevention. We also acknowledge Therese Kearns, for her assistance with data collection, and Patrick Lammie and Patricia Graves for their help interpreting the results.

